# Performance of ChatGPT as an AI-assisted decision support tool in medicine: a proof-of-concept study for interpreting symptoms and management of common cardiac conditions (AMSTELHEART-2)

**DOI:** 10.1101/2023.03.25.23285475

**Authors:** Ralf E. Harskamp, Lukas De Clercq

## Abstract

**Background:** It is thought that ChatGPT, an advanced language model developed by OpenAI, may in the future serve as an AI-assisted decision support tool in medicine.

**Objective:** To evaluate the accuracy of ChatGPT’s recommendations on medical questions related to common cardiac symptoms or conditions.

**Methods:** We tested ChatGPT’s ability to address medical questions in two ways. First, we assessed its accuracy in correctly answering cardiovascular trivia questions (n=50), based on quizzes for medical professionals. Second, we entered 20 clinical case vignettes on the ChatGPT platform and evaluated its accuracy compared to expert opinion and clinical course.

**Results:** We found that ChatGPT correctly answered 74% of the trivia questions, with slight variation in accuracy in the domains coronary artery disease (80%), pulmonary and venous thrombotic embolism (80%), atrial fibrillation (70%), heart failure (80%) and cardiovascular risk management (60%). In the case vignettes, ChatGPT’s response matched in 90% of the cases with the actual advice given. In more complex cases, where physicians (general practitioners) asked other physicians (cardiologists) for assistance or decision support, ChatGPT was correct in 50% of cases, and often provided incomplete or inappropriate recommendations when compared with expert consultation.

**Conclusions:** Our study suggests that ChatGPT has potential as an AI-assisted decision support tool in medicine, particularly for straightforward, low-complex medical questions, but further research is needed to fully evaluate its potential.

## INTRODUCTION

Artificial intelligence (AI) is considered a key asset in advancing medicine and the fundamental challenge of how to deliver sufficient healthcare for our growing and ageing populations. [1] In certain areas, such as radiology, AI is already used to make predictions and to assist in the interpretation of imaging or other diagnostic tests. [1,2] However, the true potential of AI in healthcare lies in its ability to transform clinical workflows by automating routine and time-consuming tasks. This not only improves efficiency and productivity, but also frees up healthcare providers to focus on more critical and complex tasks, where AI can serve as a valuable support tool. By combining human expertise and computational power, AI has the potential to greatly enhance the delivery of healthcare services and improve patient outcomes.[1,2]

“ChatGPT” (Chat Generative Pre-trained Transformer), an interactive large language model, is quickly gaining recognition as a potential game-changer in the field of AI-assisted decision support. [3] Since its launch as a prototype in November 2022, it has garnered significant attention (as shown in *figure 1*), with Microsoft pledging to invest billions of US dollars in this promising new technology. [4] ChatGPT also made recent headlines by passing exams from law and business schools, answering both multiple choice and essay questions. [5] Moreover, its performance at passing or nearing the passing threshold of the United States Medical Licensing Exam (USMLE) has further solidified its potential as a valuable asset in the medical field. [6] Similar models have shown a capacity for storing and reasoning with clinical information contained in their training data. [7,8] This set the pretext of this study (‘AMSTELHEART-2’), in which we explored the potential of ChatGPT as an AI-assisted decision support tool in medicine. We prompted the ChatGPT client for interpreting symptoms, in which we focused on symptoms of possible cardiac origin, where the room for error is small, with wrong judgements having important consequences. Similarly we also asked ChatGPT to help out in the management of common heart conditions.

**Figure 1.**
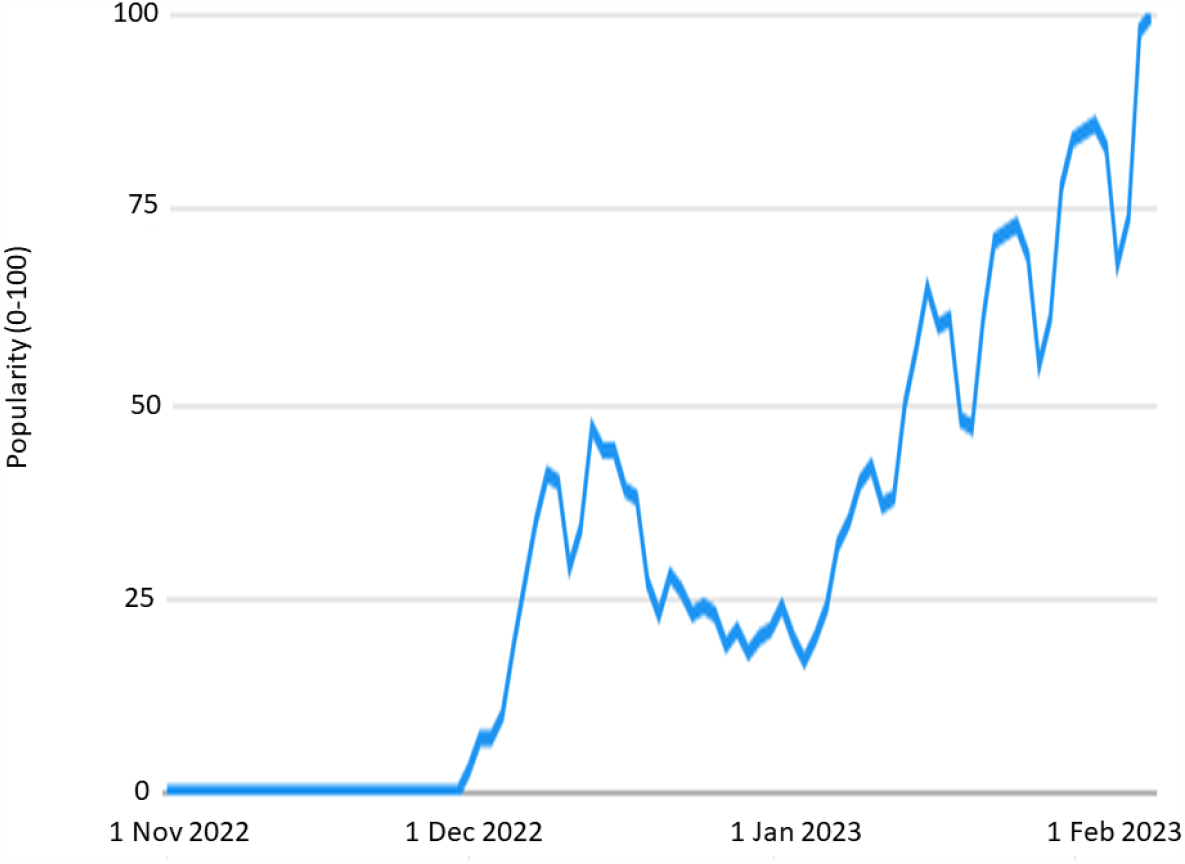
Interest over time for ChatGPT based on Google Trends Numbers represent search interest relative to the highest point on the chart for the given region and time. A value of 100 is the peak popularity for the term. A value of 50 means that the term is half as popular. A score of 0 means there was not enough data for this term. (https://trends.google.com/trends/explore?date=today%203-m&q=chatgpt) (search date: Feb 4, 2023)

## METHODS

### Design

In the AMSTELHEART-2 study, we tested the ability of ChatGPT to correctly answer cardiovascular trivia questions and to interpret symptoms or make appropriate treatment/management suggestions based on primary care based cardiovascular case vignettes. The trivia questions as well as the case vignettes were entered online on the web-based ChatGPT platform (https://chat.openai.com). We entered data in the English language only. The study was waived by the local institutional review board (W23_07#23.097).

### Index test: ChatGPT

ChatGPT is an advanced generative language model developed by OpenAI, trained on a diverse range of internet text to generate human-like responses. It uses deep learning techniques to understand and generate text, making it capable of answering questions, completing sentences, and generating text based on given prompts.[1] The prototype version of ChatGPT was launched on November 30, 2022 and for this study we relied on the Free Research Preview version of January 30, 2023.

ChatGPT is based on the Transformer architecture and utilizes deep learning techniques to generate human-like responses. The Transformer architecture was introduced in 2017 and has since become a cornerstone of modern large language models (LLMs). The architecture is designed to handle sequential data, such as text, by allowing the model to attend to different parts of the input sequence at different times, allowing it to capture long-term dependencies and contextual information. These models are trained on a large, diverse corpus of text data sourced from the internet, allowing it to learn the patterns and relationships between words and phrases. ChatGPT is a fine-tuned version of a proprietary LLM architecture developed by OpenAI. Like its sibling InstructGPT, it makes use of *Reinforcement Learning from Human Feedback*, a training technique which combines the strengths of supervised learning, reinforcement learning and human-in-the-loop approaches to improve the performance of AI models. [9,10] OpenAI continues to gather data from ChatGPT users to further train and fine-tune the model, allowing users to upvote or downvote responses and provide additional feedback through a text field. [11]

### Reference standard

For the medical trivia quizzes, the reference standard was the medical expert who developed the quiz, which was backed-up by guideline recommendations. For the case vignettes, the reference standard was the clinical advice provided by the attending physician or consulted expert and the subsequent clinical course of the corresponding patient. Two investigators also checked the advice by comparing this with guidelines recommendations.

### Test cases: trivia questions

We used Medscape’s “Fast five quizzes” for continuous medical education to gather ten questions for each of the following topics: acute coronary syndrome, pulmonary and venous thrombotic embolism, atrial fibrillation, heart failure and cardiovascular risk management. These questions can be found at: https://reference.medscape.com/index/section_10360_0

#### Test cases: case vignettes

Twenty vignettes were obtained through a random sampling of clinical cases that presented to a community health center in Amsterdam, The Netherlands. These cases were previously collected as part of the Amsterdam Heart Study, a project based on routine care data. We restricted to consultations that either involved symptoms of possible cardiac origin (chest pain, dyspnea, or palpitations), or involved questions with regards to diagnostic work-up or treatment in patients with common cardiovascular conditions. We selected 10 cases of patient-physician consultations and 10 cases of general practitioner (GP)-cardiologist/expert consultations. The case vignettes that we used were already stripped of patient identifiers via a trusted third party. In addition, we altered characteristics, such as age (we used age categories), sex, and comorbidities, so that in the end we ended up with fictionalized cases.

### Statistical methods

This study is of a descriptive nature and does not use formal statistical methodology.

## RESULTS

### Cardiovascular trivia questions

A total of 50 multiple choice questions were entered into ChatGPT’s interface. Of those questions, 37/50 (74%) were answered correctly. As shown in *Figure 2*, there was slight variation in accuracy in the domains coronary artery disease (8/10), pulmonary and venous thrombotic embolism (8/10), atrial fibrillation (7/10), heart failure (8/10) and cardiovascular risk management (6/10). Examples of incorrect statements included the duration of dual antiplatelet therapy following ACS, where ChatGPT incorrectly stated that (1) the vast majority of AF episodes are associated with significant symptoms, (2) most older patients with HF have a reduced ejection fraction, (3) an incorrect blood pressure measurement as the threshold for (stage 2) hypertension, and (4) a normal cardiopulmonary stress test with 6-minute walk test is 600 feet (approx.. 180 meters); which borders the threshold for severe cardiac dysfunction. However, ChatGPT also provided many correct answers, for instance that one should use heparin as an anticoagulant during pregnancy, on the basis of its fetal safety profile.

**Figure 2.**
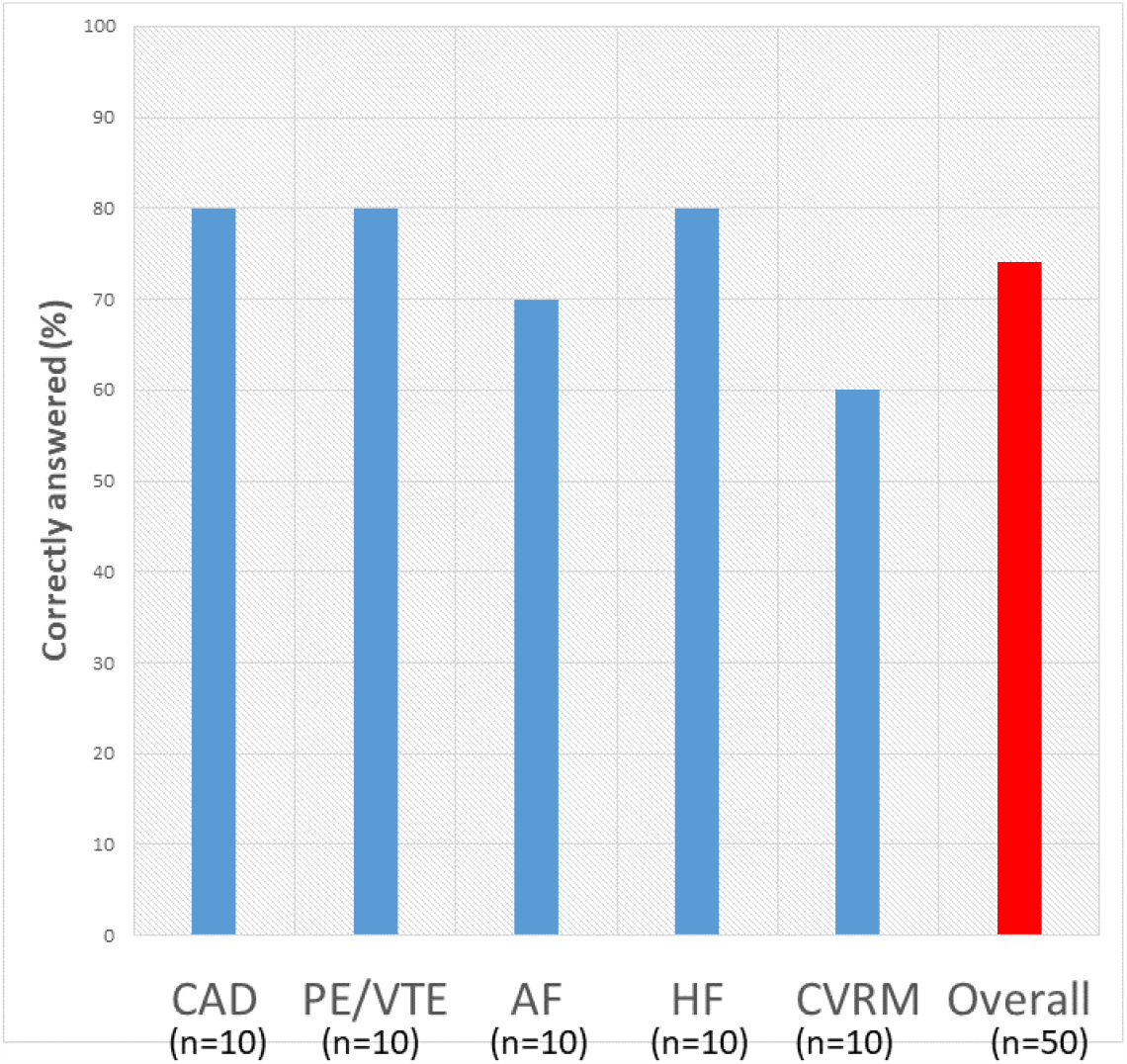
Performance of ChatGPT in correctly answering medical quizzes on key aspects and practice essentials of common cardiovascular conditions (n=50) Abbreviations: CAD= coronary artery disease (acute coronary syndrome and coronary artery atherosclerosis management); PE/VTE= pulmonary embolism and venous thromboembolism; AF=atrial fibrillation; HF= heart failure; CVRM= cardiovascular risk factors and management.

### Case vignettes regarding patients reaching out to primary care with a cardiac complaint

*Table 1* displays the characteristics of 10 patients and their questions for which they reached out to their primary care provider. Questions were related to whether symptoms should be a reason for concern and for consulting their GP, questions about medication use, as well as on behavioral changes, ranging from diet to resuming sexual activity after a myocardial infarction. Overall, ChatGPT provided advice that was in line with the actual advice given and care provided (9/10). We noted one major inconsistency, in which ChatGPT advised to use of thrombolytic agents as secondary preventative medications in a patients with a prior myocardial infarction (MI). While there is a place for thrombolytics in the acute phase (in settings where primary PCI is not available), these medications have no place in chronic management of patients post MI.

**Table 1.**
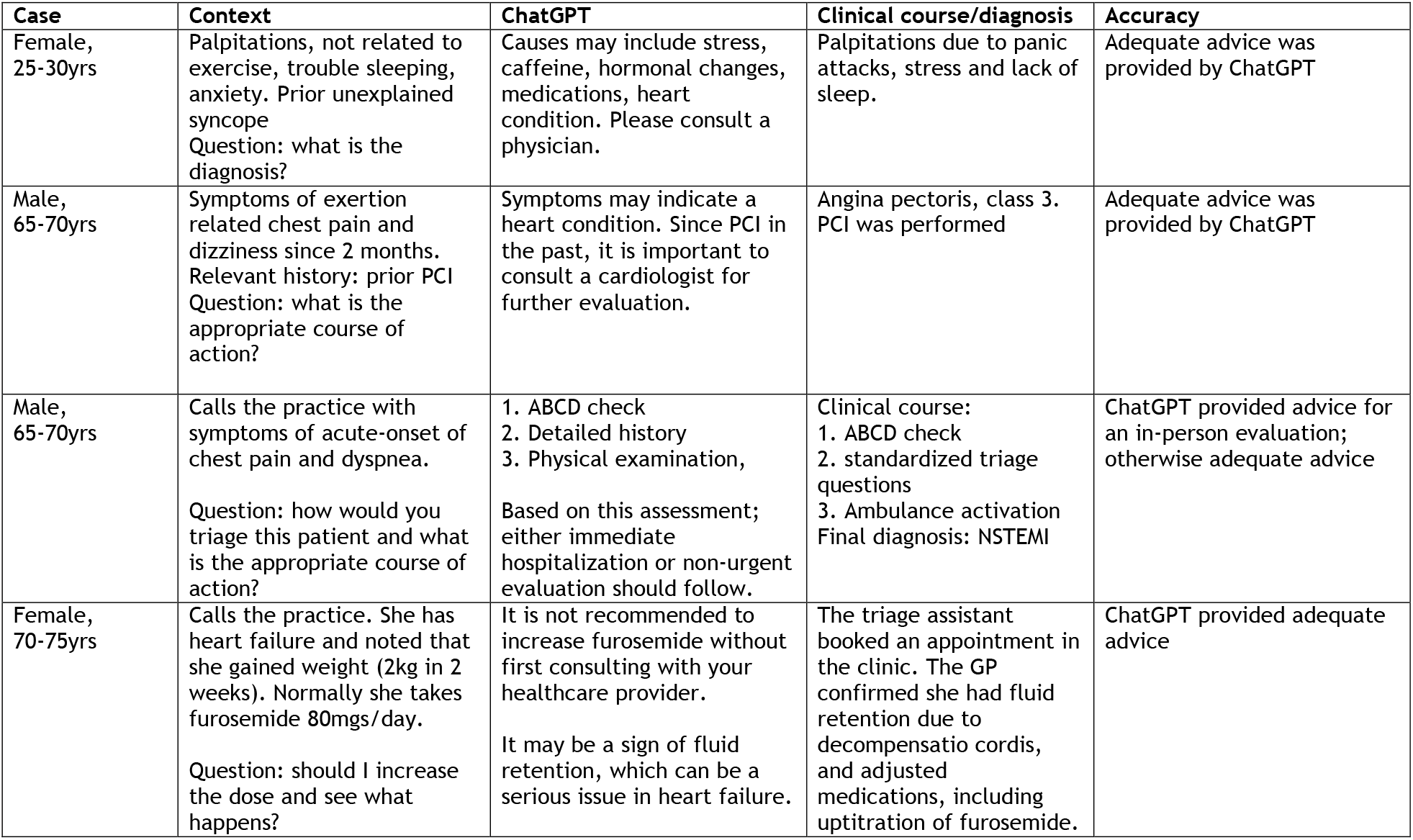
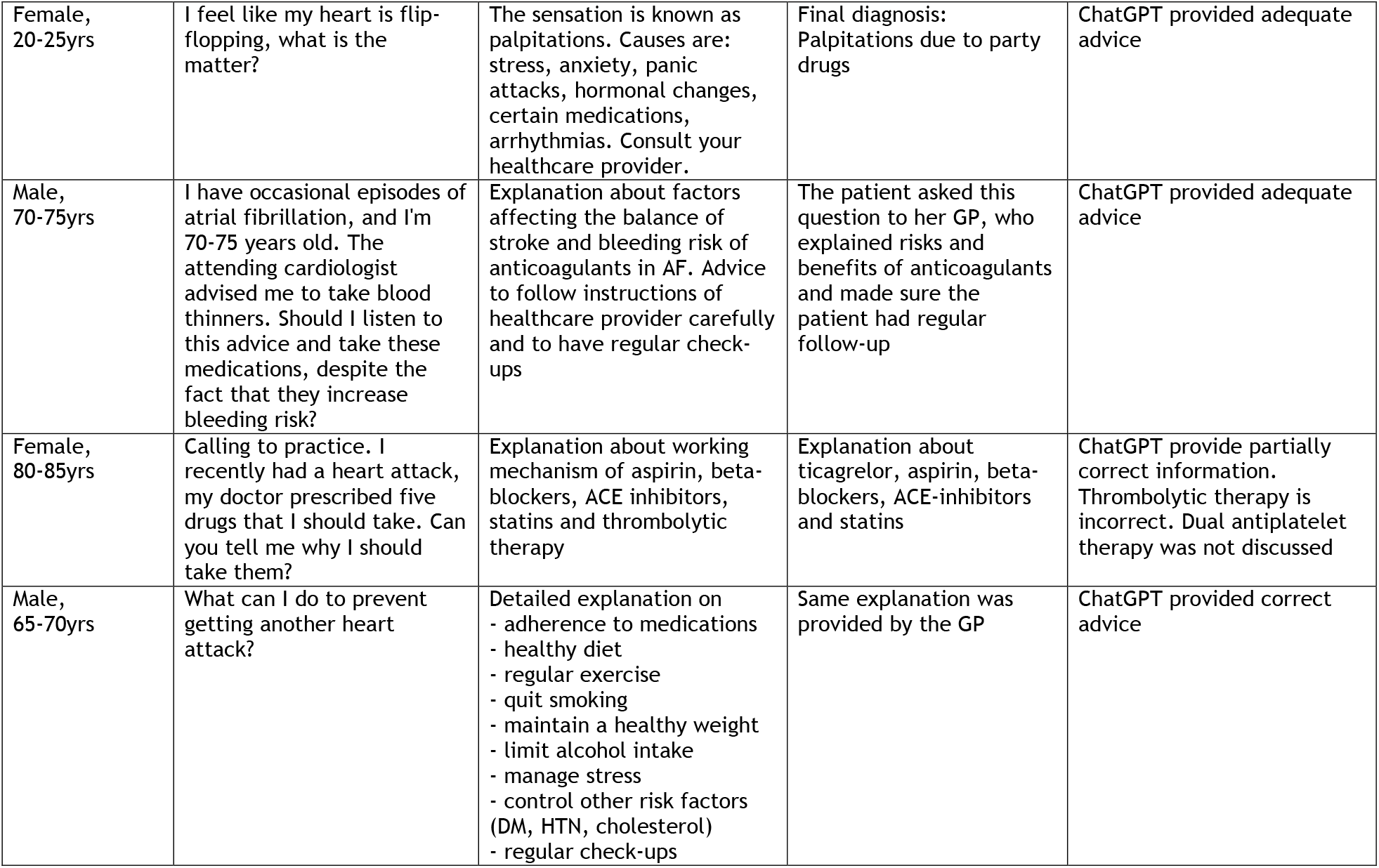
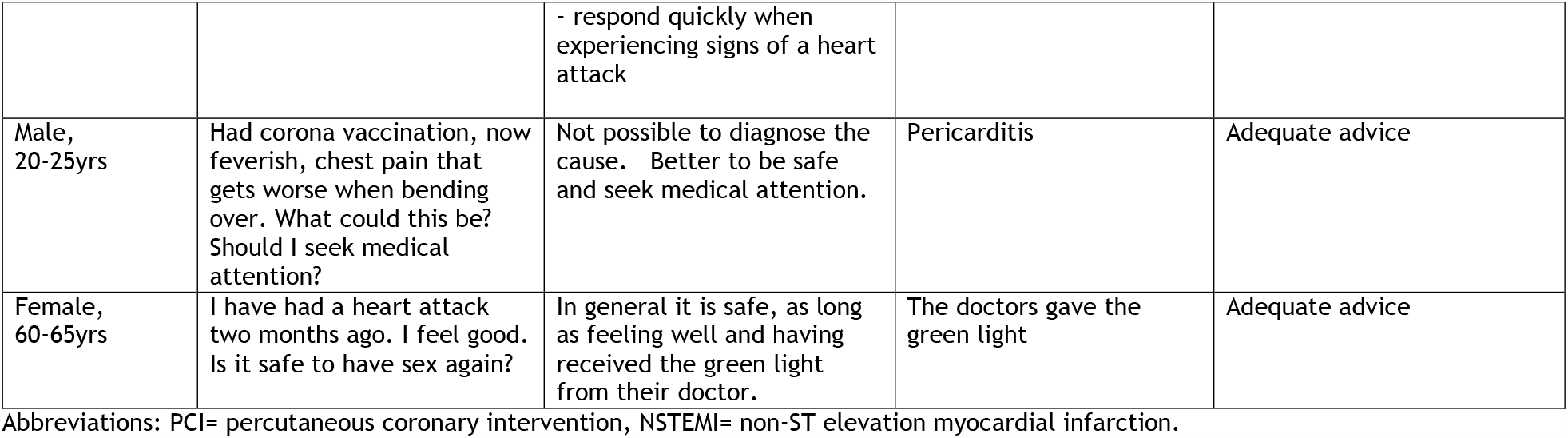
ChatGPT recommendations based on medical questions from patients and accuracy when compared with actual clinical course by a healthcare provider

### Case vignettes regarding physicians asking for expert consultation

*Table 2* displays the 10 questions that GPs send out for expert consultation, either to a cardiologist or to a GP with special interest in cardiology via digital consultation. We also fed these questions to ChatGPT, and found that out of the 10 questions, five answers (50%) fully matched that of the advice provided by the expert. Two answers (20%) were a partial match, one question (10%) was inconclusive, and two answers were incorrect (20%).

**Table 2.**
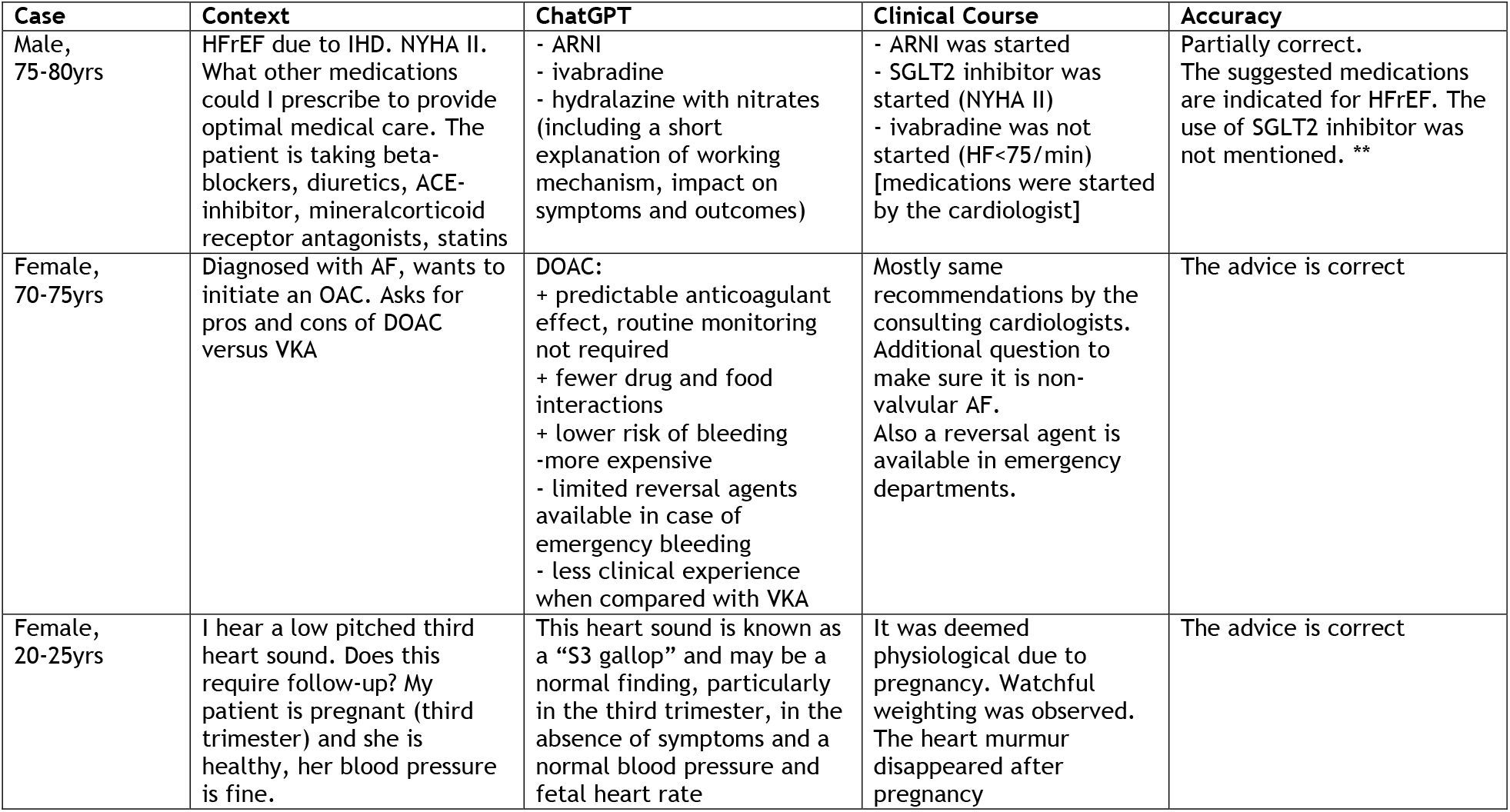
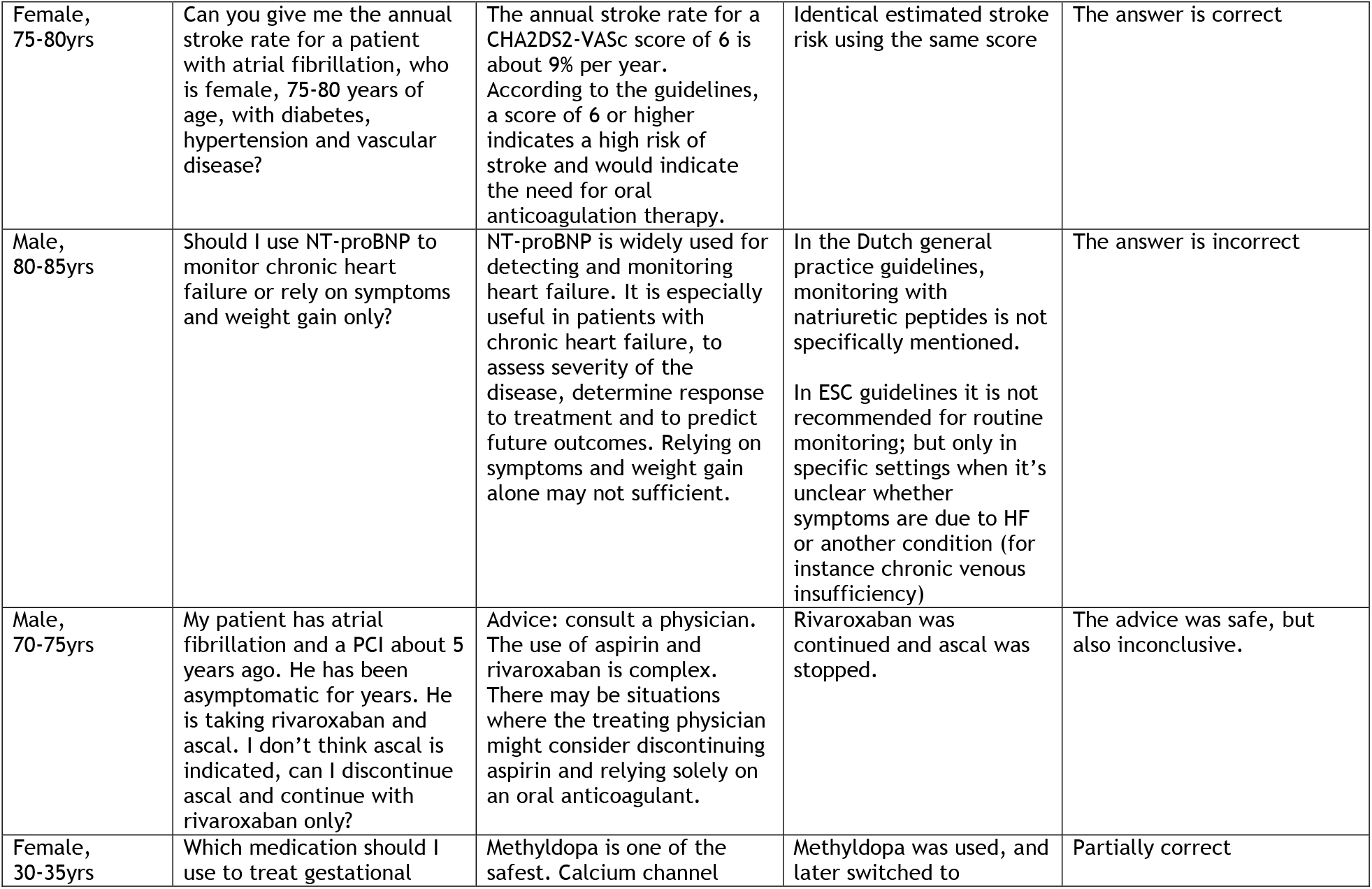
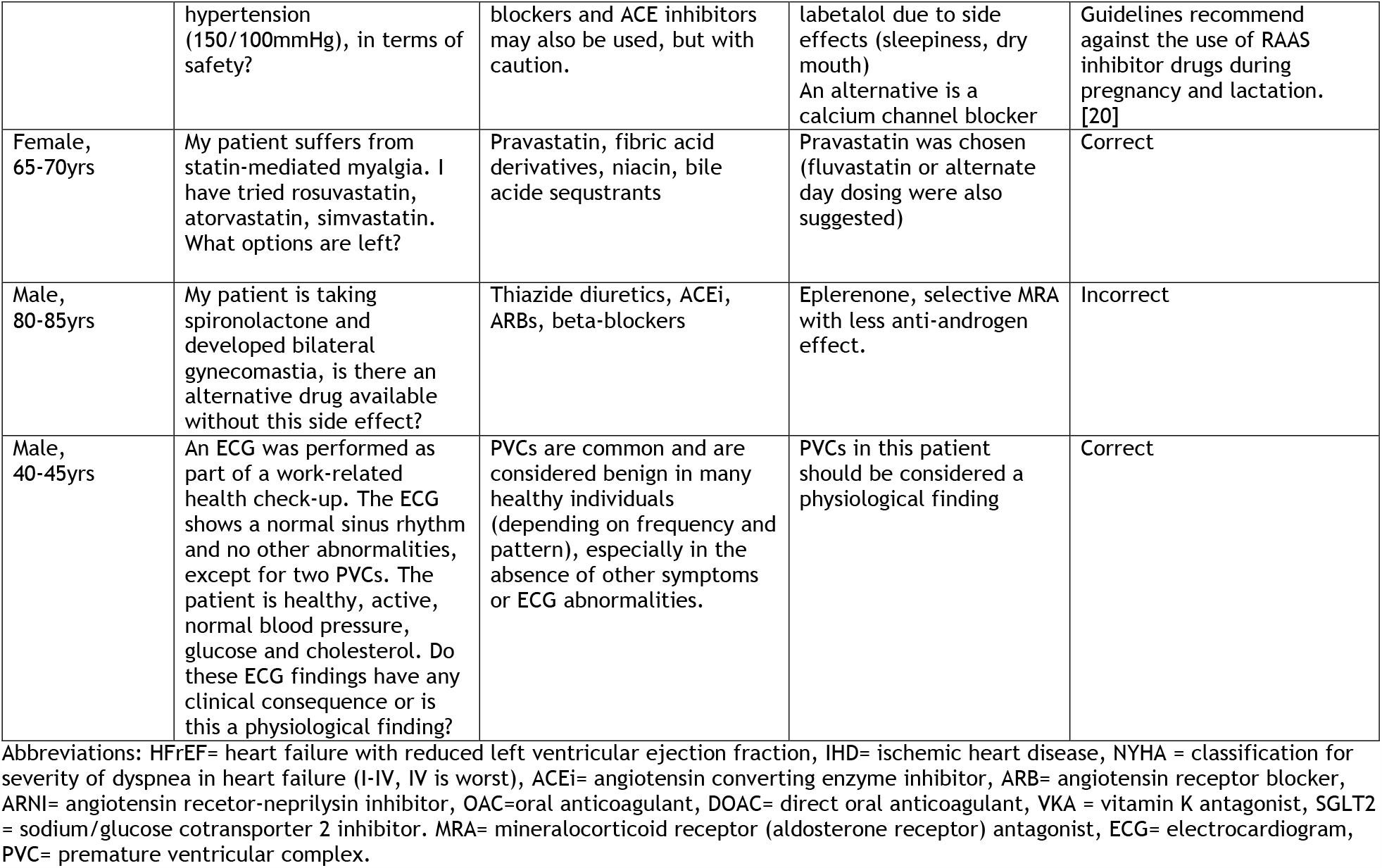
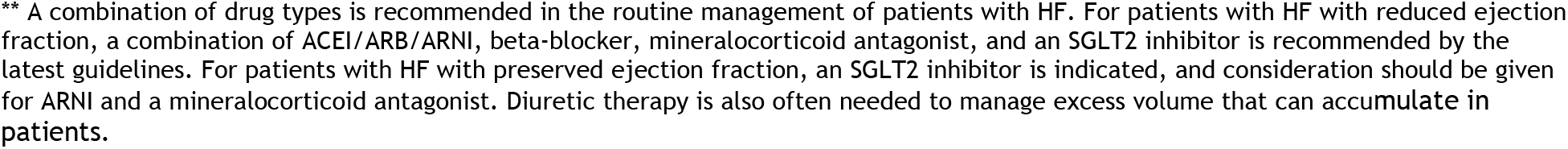
ChatGPT recommendations based on medical questions from physicians and accuracy when compared with actual clinical course by collegial consultation

## DISCUSSION

In the AMSTELHEART-2 study we investigated the feasibility of using a generative language model, ChatGPT, in the interpretation of cardiac symptoms and the provision of preliminary diagnosis suggestions. Our hypothesis was that the model’s ability to process and comprehend natural language input and capacity for storing clinical information would enable it to extract relevant information and generate potential diagnoses based on the reported symptoms. The results showed that multiple choice questions part of a continuing cardiovascular education course were overall sufficiently answered (accuracy of 74%). Moreover, relative straightforward medical questions coming from patients reaching out to primary care could be well addressed in the vast majority of cases.

However, our exploratory analysis also revealed that ChatGPT was insufficient in addressing medical questions from physicians who consulted their expert-peers. This suggests that ChatGPT was not sufficiently trained with this type of data, and may not have fully comprehended the nuanced context of the medical questions that we provided it with.

### Strengths and limitations

A strength of this study is that we relied on multiple approaches (trivia questions and case vignettes with variable complexity) to evaluate ChatGPT’s accuracy in addressing medical issues. Particularly the use of the cases vignettes provides a good simulation of how ChatGPT would perform in real-life. A limitation of the study is the relative small sample size; however despite the limited number of observations, we were able to see patterns in the variable performance of ChatGPT. A different study design, such as a head-to-head comparison between an AI-assisted triage tool in primary care versus usual care, would perhaps have been even more informative. However, given the early stage of the technology, we believe it’s premature to consider such an approach, and further refinements should first be made.

### Prior studies

At the time of writing a total of 20 medical studies were published on ChatGPT. Most of these studies underline the promise of ChatGPT. For instance, there are reports on how it stirs (medical) education, as it can generate convincing abstracts and essays, or write computer code. [12,13] There are also projects that use AI models to generate medical reports, for example for radiology, helping radiologists to quickly and accurately interpret and report the results of medical imaging. To stir up things more, researchers found that ChatGPT performed reasonably well on the US medical licensing exam. [6] This opens up opportunities to explore how well it would do when addressing medical questions in real-life. Data on this is scant, and the findings from our study may present one of the first efforts in the field of community-based medicine.

### Future perspectives

Not that long ago, primary care based physicians considered the potential of artificial intelligence to be limited. [14] However, this view will likely change dramatically this decade, given the enormous interest of the community for this new technology. So what could these AI models do to advance medicine? First of they can process large amounts of data quickly and accurately, and can identify patterns and relationships in data that would be difficult for humans to detect on their own. Technology, such as ChatGPT could aid doctors and patients in a various ways. One application is that the technology may quickly and accurately identify possible illnesses, based on the patient’s symptoms and medical records. When patients have direct access to this technology it could lead them to seek medical care in a timely manner, which in turn leads to faster and more accurate diagnoses and treatment plans. From a physician’s perspective, ChatGPT could be used to advise on medical decisions, thanks to its ability to analyze large volumes of medical data to identify patterns that influence diagnostic or treatment decisions. This allows to make better-informed and more accurate decisions when they are treating patients.

From a research perspective, ChatGPT could also be a major help in the development and testing of new drugs, treatments, and medical technologies. By analysing data from past trials and experiments, ChatGPT could better optimise the process and allow for faster, more informed decisions on new drug development and treatments. This could bring about improvements in the speed, efficacy, and cost-effectiveness of medical developments.

ChatGPT may also help in writing down new scientific discoveries and findings. It is though that ChatGPT will shape the future of medical writing, with human judgment and oversight for quality assurances. [15,16] Already, some authors have gone as far as crediting ChatGPT with formal authorship [17]. In response, the prestigious journal Nature has made a recent statement that no large language models will be accepted as credited author on a research paper in their journal (or any journal of the Springer group), for the simple reason that AI tools cannot take accountability for the work. [17]

### Concerns

ChatGPT has shown limitations akin to those found in preceding LLMs. Like its predecessors, it is not immune to biases. Models such as these are trained on large datasets that are sourced from various websites, books, and other sources, some of which may contain implicit or explicit biases. Despite attempts to address this issue, there have been numerous instances where the model has exhibited biases related to race, gender, politics, and other areas. [18] Whether this behaviour is maintained for medical topics remains to be seen, but can be expected.

Being a probabilistic language model, ChatGPT may generate different outcomes for identical inputs. This hinders evaluation of its performance, identification of biases, and can be a concern in medical applications where accuracy and consistency are critical. The development and implementation of such non-deterministic language models in healthcare should involve collaboration between experts in fields such as medicine, computer science, ethics, and law.

Training LLMs requires vast amounts of computational resources and energy, leading to high financial costs which present a barrier to entry for smaller organizations or researchers. Development of these models has been limited to a handful of large companies with extensive resources, many of which do not publish their full models or details regarding their training, including OpenAI. [19] The selective access to these models and non-transparency of their training can lead to a concentration of power and control over AI technology in the hands of a few companies, which can have negative implications for competition, innovation, and social responsibility in AI. It is important to strive for greater transparency in the training and fine-tuning of LLMs like ChatGPT to ensure responsible and trustworthy development and use of AI in medicine.

## CONCLUSION

In this study we explored whether ChatGPT, a large language model, can be used as an AI-support tool in medicine, by asking it to interpret symptoms and provide initial suggestions for diagnosis and management. We found it performed well when posed multiple-choice questions, as well as when it involved more straightforward questions from patients reaching out to their primary care physician. The model had more difficulty answering more advanced medical questions. Multidisciplinary research is needed to figure out what the right place for ChatGPT could be in a community that is quickly starting to embrace this new form of technology. It is important to consider the risks associated with the non-determinism, non-transparency, and increased centralization of LLMs like ChatGPT and strive for greater accountability in their development and use, particularly in the critical and sensitive context of healthcare.

## Data Availability

All data produced in the present study are available upon reasonable request to the authors

## REFERENCES

[1] Aung YYM, Wong DCS, Ting DSW. The promise of artificial intelligence: a review of the opportunities and challenges of artificial intelligence in healthcare. British Medical Bulletin 2021;139:1:4–15.

[2] Marr B. How AI and machine learning will impact the future of healthcare. Forbes. https://www.forbes.com/sites/bernardmarr/2022/09/14/how-ai-and-machine-learning-will-impact-the-future-of-healthcare/?sh=5fa512bc47e5

[3] ChatGPT, OpenAI. Available at: https://openai.com/products/gpt-3/ accessed on January 29 – February 2nd, 2023.

[4] No authors. Microsoft and OpenAI extend partnership. https://blogs.microsoft.com/blog/2023/01/23/microsoftandopenaiextendpartnership/

[5] Murphy Kelly S. ChatGPT passes exams from law and business schools. https://edition.cnn.com/2023/01/26/tech/chatgpt-passes-exams/index.html

[6] Kung TH, Cheatham M, ChatGPT, et al. Performance of ChatGPT on USMLE: potential for AI-assisted medical education using large language models. https://www.medrxiv.org/content/10.1101/2022.12.19.22283643v1.full

[7] Singhal K, Azizi S, Tu Tao, et al. Large language models encode clinical knowledge. https://arxiv.org/pdf/2212.13138.pdf

[8] Liévin V, Egeberg Hother C, Winther O. Can large language models reason about medical questions? https://arxiv.org/pdf/2207.08143.pdf

[9] Ouyang L, Wu J, Jiang X, et al. Training language models to follow instructions with human feedback. https://arxiv.org/pdf/2203.02155.pdf

[10] Stiennon N, Ouyang L, Wu J, et al. Learning to summarize from human feedback. https://arxiv.org/pdf/2009.01325.pdf

[11] No authors. ChatGPT Feedback Contest: Official Rules. https://cdn.openai.com/chatgpt/ChatGPT_Feedback_Contest_Rules.pdf

[12] Else H. Abstracts written by ChatGPT fool scientists. Nature 2023;613:423.

[13] Looi Mun-Keat. Sixty seconds on… ChatGPT. BMJ 2023; 380:205.

[14] Blease CR, Bernstein M, Kaptchuk TJ, Mandl KD. Artificial intelligence and the future of primary care: an exploratory qualitative study of UK GPs’ Views. J Med Inter Res 2018; DOI: 10.2196/12802.

[15] Kitamura FC. ChatGPT is shaping the future of medical writing but still requires human judgment. Radiology 2023;2:230171.

[16] Biswas S. ChatGPT and the future of medical writing. Radiology 2023;2:223312.

[17] No authors listed. Tools such as ChatGPT threaten transparent science; here are our ground rules for their use. Nature 2023;613:612.

[18] Zhuo TY, Huang Y, Chen C, Xing Z. Exploring AI ethics of ChatGPT: a diagnostic analysis. https://arxiv.org/pdf/2301.12867.pdf

[19] Goldstein JA, Sastry G, Musser M, DiResta R, Gentzel M, Sedova K. Generative language models and automated influence operations: emerging threats and potential mitigations. https://arxiv.org/pdf/2301.04246.pdf

[20] Williams B, Mancia G, Spiering W, Agabiti Rosei E, Azizi M, Burnier M, Clement DL, et al; ESC Scientific Document Group. 2018 ESC/ESH Guidelines for the management of arterial hypertension. Eur Heart J. 2018;39:3021–104.

